# Integrative, and Scalable mental health phenotyping using a knowledge-graph-derived dual-metric framework

**DOI:** 10.64898/2026.03.09.26347798

**Authors:** Arushi Sharma, Akshay Bharadwaj, Shivani Modi, Gaurav Ahuja, Ankur Jain, Kaushal Kumar

## Abstract

Prevailing diagnostic instruments for anxiety and depression, though clinically indispensable, remain anchored to symptom-focused queries that assess patients directly about their affective states, while often neglecting the multidimensional architecture of daily living. Here, we introduce two complementary metrics, the Cognitive Attention Score (CAS) and C:ERR (Cognition-to-Emotional-Response Ratio), derived from yogic psychology and operationalized within a structured knowledge graph (Ceekr-KG) comprising 151,288 triples linking 354 discrete CAS levels, 26 continuous C:ERR values, and 80 clinical symptoms. Rather than interrogating disease phenotypes directly, these metrics are computed by capturing circadian, nutritional, and lifestyle factors that jointly regulate cognitive and emotional homeostasis. Hyperparameter-tuned Ceekr-KG model demonstrated high structural fidelity (Hits@1 = 97%, mean reciprocal rank = 0.98), substantially outperforming relation-preserving randomized controls, indicating that predictive performance arises from semantic structure rather than graph topology alone. CAS and C:ERR showed a strong positive association (Spearman’s ρ = 0.787, p < 0.0001) but exhibited distinct distributional properties, with C:ERR displaying consistently stronger inverse correlations with symptom severity across domains (e.g., low energy: ρ = -0.85 versus -0.70 for CAS). Ordinal regression further showed that a combined CAS and C:ERR model outperformed either metric alone for most symptoms, indicating complementary and non-redundant contributions to clinical variance. Integration of Ceekr-KG into the independent Clinical Knowledge Graph improved predictive performance of widely used questionnaire-based assessment scales, demonstrating that yogic psychological frameworks encode clinically relevant semantic information. Finally, longitudinal analysis of 249 individuals meeting predefined inclusion criteria (baseline CAS < 64 and >=2 assessments) across three therapeutic programmes revealed a mean CAS increase of +11.45 points (p < 0.001) and substantial migration from lower to higher functional bands, establishing Ceekr-KG as a validated digital phenotype for scalable mental health assessment.

## Introduction

Mental health disorders represent one of the most pressing global health challenges of the twenty-first century. Anxiety and depressive disorders affect over 300 million and 280 million people worldwide, respectively, constituting the leading cause of disability and incurring economic losses exceeding USD 1 trillion annually ^[1,2]^. Despite this growing burden, accurate, scalable, and holistic assessment of mental health remains an elusive goal. Contemporary mental health diagnostics rely heavily on clinician-administered scales such as the Hamilton Anxiety Rating Scale (HAM-A) ^[3]^, Hamilton Depression Rating Scale (HAM-D) ^[4]^, and Generalized Anxiety Disorder-7 (GAD-7) ^[5]^. While these scales possess established psychometric properties, they carry inherent limitations that constrain their utility in modern healthcare. First, they are time-intensive, requiring 20-25 minutes of clinician time per administration, rendering them impractical for large-scale screening or frequent longitudinal monitoring. Second, reliance on expert interpretation introduces inter-rater variability and limits accessibility in underserved regions where mental health professionals are scarce. Third, these scales assess anxiety and depression separately despite their well-documented comorbidity, potentially overlooking the integrated nature of mental health. Fourth, they capture only cognitive and emotional symptoms while neglecting the intricate interplay between mental states, lifestyle factors, and physiological processes that contemporary research has shown to be fundamental to mental well-being ^[6]^.

Beyond questionnaire-based methods, invasive techniques such as neuroimaging, electroencephalography, and biomarker assays have been explored for mental health assessment ^[7,8]^. Functional magnetic resonance imaging (fMRI) has identified aberrant connectivity patterns in depression and anxiety ^[9]^, while electroencephalographic markers such as frontal alpha asymmetry have shown promise as correlates of affective style ^[10]^. Peripheral biomarkers, including inflammatory cytokines and cortisol, have been extensively investigated as potential objective indicators ^[11]^. However, these approaches face substantial barriers to clinical translation: high cost, limited accessibility, poor specificity, and lack of standardised protocols preclude their deployment as routine screening tools. Moreover, such reductionist approaches fail to capture the phenomenological richness essential for personalised care. Parallel to these biomedical developments, ancient contemplative traditions have long offered holistic frameworks for understanding mental well-being. The yogic psychology traditions of the Indian subcontinent provide sophisticated phenomenological taxonomies of mind, attention, and consciousness refined over millennia ^[12–14]^. Ayurvedic medicine emphasises links between diet, sleep, routine, and mental health that modern psychiatry is only beginning to systematically investigate ^[15,16]^. Despite the sophistication of these frameworks, they have remained outside evidence-based practice due to the absence of rigorous operationalisation and empirical validation. The emergence of knowledge graphs (KGs) as a powerful computational paradigm for integrating heterogeneous biomedical data offers a unique opportunity to bridge this ancient-modern divide. Knowledge graphs represent entities as nodes and their relationships as edges, enabling sophisticated reasoning and discovery across interconnected datasets. The Clinical Knowledge Graph (CKG) integrates over 20 million nodes and ∼700 million relationships from experimental data, public databases, and literature, demonstrating the power of graph-based approaches to capture the multidimensional complexity of human health ^[17]^. However, the application of KGs to mental health assessment, particularly for validating diagnostic scales, remains largely unexplored.

Here, we introduce two complementary metrics derived from yogic psychology and translated into dual quantitative scores, i.e., the Cognitive Attention Score (CAS) and C:ERR (Cognition-to-Emotional-Response Ratio). Operationalised within the structured knowledge graph Ceekr□KG, these metrics quantify cognitive-attentional state and cumulative emotional dysregulation using self-reported information on sleep, nutrition, and exercise. Ceekr□KG was constructed from 151,288 triples linking 354 CAS levels, 26 continuous C:ERR values, and 80 clinical symptoms. We validated its structural integrity using state-of-the-art embedding models, benchmarked CAS against gold-standard test (HAM-A, HAM-D, GAD-7) via the independent Clinical Knowledge Graph, and analysed longitudinal trajectories from a cohort of 1,840 individuals, of whom 249 met predefined inclusion criteria (baseline CAS < 64; ≥2 longitudinal assessments) across three therapeutic programmes. Integrating Ceekr□KG into CKG systematically improved predictive performance across all scales, demonstrating that our framework encodes novel, clinically relevant knowledge. By translating ancient contemplative wisdom into a validated AI-driven computational framework, this work establishes Ceekr□KG as a robust, scalable digital phenotype for precision mental health.

## Results

### CAS and C:ERR exhibit distinct distributions, strong correlation, and complementary associations with clinical symptoms

The Cognitive Attention Score (CAS) and the Cognition-to-Emotional-Response Ratio (C:ERR) are derived from a structured knowledge matrix that formalizes yogic psychology principles into a computational framework linking both metrics to 80 clinical symptoms across physical, cognitive, and emotional domains. Although complementary, the two indices quantify distinct aspects of mental health: CAS reflects cognitive attentional state based on self-reported sleep, nutrition, and physical activity, whereas C:ERR represents a weighted composite of cumulative cognitive and emotional dysregulation **(Fig. 1A)**. Following post-processing of the assimilated data derived from yogic psychology principles, the dataset was converted into a machine-readable matrix comprising 1,073 rows and 82 columns, jointly representing CAS, C:ERR, and 80 symptom variables. CAS ranged from 22.06 to 107.89, and C:ERR from 0.2 to 2.7 **(Supplementary Fig. 1A)**. We first examined their distributions. CAS showed a clearly multimodal profile with density peaks near ∼30, ∼60, and ∼85 **(Fig. 1B, upper; Supplementary Table 3)**. In contrast, C:ERR displayed a right-skewed distribution with a dominant mode near 0.9 and a long tail extending to ∼2.5, with most values between 0.5 and 1.2 **(Fig. 1B, lower)**. Quantile-Quantile plots and Shapiro-Wilk tests confirmed significant non-normality for both metrics (p < 0.001; **Fig. 1C**), motivating non-parametric analyses. Despite their distinct marginal distributions, CAS and C:ERR were strongly correlated **(Fig. 1D, Supplementary Fig. 1D)**.

**Figure 1:**
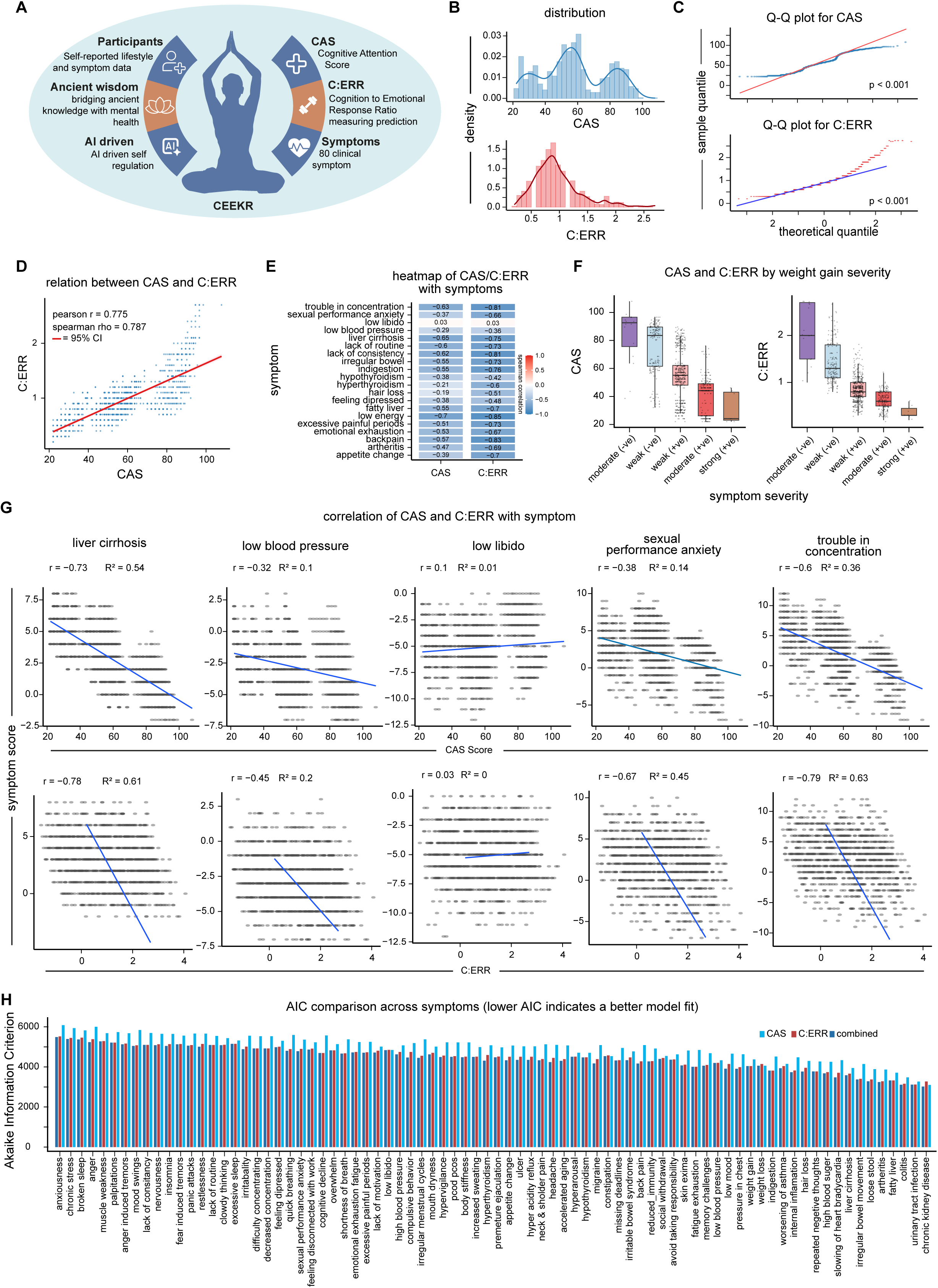
Distributional properties, inter-relationship, and clinical associations of CAS and C:ERR. **(A)** Schematic overview of the full study cohort and computational framework linking participants, CAS, C:ERR, and clinical symptoms. **(B)** Distribution of Clinical Assessment Scores (CAS) and Error-Corrected C-scores (C:ERR). Histograms represent empirical density frequencies overlaid with Gaussian kernel density estimates. (Upper panel) CAS distribution across the study population, exhibiting a multi-modal pattern with distinct peaks at approximately 30, 60, and 85. These peaks suggest three latent subpopulations: low scorers (severe impairment), average scorers (moderate impairment), and high scorers (minimal impairment). (Lower panel) C:ERR distribution, displaying a primary mode near 0.9 with a distinct right-skew (positive skewness). While most values are concentrated between 0.5 and 1.2, a long tail extends to 2.5, indicating a subset of participants with higher error rates. **(C)** Quantile-Quantile (Q-Q) plots comparing CAS and C:ERR distributions against theoretical normal quantiles. Red and blue solid lines represent the expected path of a perfectly normal distribution. Shapiro-Wilk test confirmed significant non-normality for both metrics (p < 0.001), validating the use of non-parametric methods. **(D)** Scatter plot illustrating the relationship between CAS and C:ERR. The red line represents a linear regression fit with 95% confidence interval shaded in pink. Pearson’s r = 0.775 (p < 0.001), Spearman’s ρ = 0.787 (p < 0.001), indicating robust positive association with approximately 60% shared variance (R² ≈ 0.60). **(E)** Correlation heatmap of CAS and C:ERR with the 20 most frequently reported clinical symptoms, selected based on data density (frequency of non-missing responses). Heatmap displays Spearman rank correlation coefficients (ρ). Color gradient: dark blue = strong negative correlation, white = no correlation, red = strong positive correlation. Numerical values within each tile provide specific correlation coefficients. C:ERR consistently exhibits stronger correlations (darker blue) than CAS across most symptoms, indicating superior sensitivity to symptomatic distress. Low libido shows near-zero correlation with both metrics (ρ ≈ 0.03). **(F)** Boxplots showing distribution of CAS (left) and C:ERR (right) across five severity levels of weight gain (negative, weak negative, weak positive, positive, strong positive). Boxes represent median and interquartile range (IQR); whiskers denote range; jittered points overlay individual participant data. Both metrics exhibit step-wise monotonic decline with increasing severity, with C:ERR showing more pronounced decline, particularly at the transition from negative to weak negative. At strong positive severity, data points cluster tightly at the bottom of the scale, confirming severe weight gain as a reliable predictor of low scores. **(G)** Scatter plots showing the relationship between CAS and CERR scores with top symptoms. Each point represents an individual participant. Blue lines indicate linear regression fits. Pearson correlation coefficients (r) and coefficients of determination (R²) are displayed within each panel. **(H)** This grouped horizontal bar chart compares the fit of three models for each specific symptom. Shorter bars (lower AIC values) indicate a better statistical fit. The color coding distinguishes between CAS (skyblue), C:ERR (red), and combined (darkblue) models.

Pearson’s r was 0.775 (p < 0.001; R² ≈ 0.60), and Spearman’s ρ was 0.787 (p < 0.001), indicating a robust monotonic relationship across the full range of values.

We next assessed associations with symptom severity across 80 symptoms, focusing on the 20 with most prominent associations **(Fig. 1E)** Both metrics showed pervasive negative correlations, with C:ERR consistently exhibiting larger effect sizes. The strongest associations were observed for fatigue/exhaustion/low energy (ρ = -0.85 for C:ERR vs -0.70 for CAS), back pain (-0.83 vs -0.57), trouble in concentrating (-0.81 vs -0.63), lack of consistency (-0.81 vs - 0.62), and cognitive decline (-0.78 vs -0.76). Low libido showed near-zero correlation with both metrics (ρ ≈ 0.03), indicating statistical independence. To further illustrate these relationships, we next visualized representative symptom-score relationships **(Fig. 1G)**. Across selected symptoms, negative associations were again evident, with steeper regression slopes observed for C:ERR relative to CAS. Cognitive symptoms such as trouble in concentration demonstrated particularly strong inverse relationships, whereas low libido remained largely uncorrelated, reinforcing its relative independence from systemic score variation. Comparative boxplots of CAS across top clinical indicators shows the distribution of CAS scores across severity levels (strong negative to strong positive) for the most frequently reported symptoms. Feeling depressed shows a sharper drop between weak-positive and positive levels, suggesting a threshold effect in which the systemic score collapses once a certain severity is reached. Fatigue emerges as one of the most reliable predictors of low CAS, evidenced by clear, non-overlapping medians across severity categories **(Supplementary Fig. 1B)**. Using weight gain as an example, both CAS and C:ERR declined monotonically across five ordered severity levels **(Fig. 1F)**. The decrease was steeper for C:ERR, particularly from moderate negative to weak negative severity. At the highest severity level, values for both metrics clustered near the lower bound, indicating that severe symptoms reliably predict low CAS and C:ERR. We then fitted ordinal logistic regression models predicting symptom severity using CAS alone, C:ERR alone, or both combined, and compared models using AIC **(Fig. 1H; Supplementary Fig. 1C)**. The combined model achieved the lowest AIC for 71 of 80 symptoms, whereas C:ERR alone was optimal for 8 and CAS alone for only 1. Across the symptom spectrum, combined and C:ERR models consistently achieved lower AIC values than CAS alone, visually confirming that CAS provides the weakest relative predictive fit. Symptoms positioned at the bottom of the distribution, such as worsening of asthma and avoiding taking responsibility, exhibited the lowest overall AIC scores, indicating they are more “predictable” within this framework than symptoms like anxiousness. While cognitive decline showed near-equivalent explanatory power between the C:ERR and combined models ΔAIC < 2), other symptoms relied on the non-redundant information provided by the combined approach. This stratification demonstrates that symptom predictability is jointly modulated by specific relational semantics and disease severity. Joint analysis of Spearman correlations and AIC values further stratified symptom classes **(Supplementary Fig. 1E)**. Colitis, fatty liver, and urinary tract infection showed both the strongest negative correlations with C:ERR and the lowest AIC values, identifying them as core symptoms most reliably predicted by C:ERR. Low libido localized to the high-AIC region, confirming independence from C:ERR, whereas anxiousness and chronic stress showed greater dispersion, consistent with higher variability in psychological symptom expression. Hierarchical clustering revealed coherent symptom groups with shared severity profiles **(Supplementary Fig. 4A)**. Together, these results establish CAS and C:ERR as complementary, strongly correlated but non-redundant metrics that stratify symptom burden, with C:ERR capturing core somatic-cognitive severity and CAS providing critical additional explanatory power for specific clinical phenotypes.

### Translating the CAS/C:ERR matrix into a predictive knowledge graph

The knowledge matrix underlying the Cognitive Attention Score (CAS) and the Cognition-to-Emotional-Response Ratio (C:ERR), which encodes their quantitative relationships to 80 clinical symptoms, was translated into a knowledge graph to enable graph-based predictive modeling. By instantiating these relationships as subject-predicate-object triples, we constructed the Ceekr Knowledge Graph (Ceekr-KG), yielding a graph with 460 nodes and 151,288 triples across six edge types encoding incremental and decremental relations **(Fig. 2A)**. The graph comprised 354 discrete CAS levels, 26 C:ERR levels, and 80 symptom nodes, with edge types capturing both the polarity and strength of association **(Fig. 2B, C)**. We next trained multiple knowledge-graph embedding models, including Diagonal Matrix (DisMult) (a bilinear diagonal embedding model)^[18,19]^, RotatE (rotational embeddings in complex space)^[20]^, ComplEx (complex-valued embeddings)^[21]^, RESCAL (relational scalable latent factorization^[22]^), and SimplE (simple embeddings based on canonical polyadic decomposition)^[23]^, under identical base parameters (batch size 1,024, embedding dimension 16, learning rate 0.01, 1182 training steps; **Fig. 2K, Supplementary Fig. 5H)** and evaluated performance using standard link-prediction metrics (Hits@1, Hits@3, Hits@10, and mean reciprocal rank) **(Supplementary Fig. 5E)**. Among all base models, SimplE consistently achieved the highest performance on both test and validation sets **(Fig. 2D,E)**. On the test set, SimplE achieved Hits@1 = 82.7%, Hits@3 = 97%, Hits@10 = 100%, and MRR = 0.90, and demonstrated the fastest and smoothest convergence during training **(Fig. 2F)**. To further improve predictive performance, we performed systematic hyperparameter optimization of the SimplE model **(Fig. 2K)**. The hypertuned SimplE model increased embedding dimension (128), gamma (21), learning rate (0.5), and maximum training steps (2,500). This optimization resulted in substantial performance gains **(Fig. 2G)**, with Hits@1 improving from 82.7% to 96.9%, Hits@3 from 97.4% to 99.8%, and MRR from 0.90 to 0.98, while maintaining perfect Hits@10. These results demonstrate that performance improvements arise not only from model choice but also from optimized training dynamics. The model’s robustness was confirmed by 5-fold cross-validation, which yielded highly consistent performance (Hits@1 = 95-97%, Hits@3 = ∼99%, mean reciprocal rank =∼97%; **Fig. 2H**). To determine whether this performance reflected genuine semantic structure rather than graph topology, we evaluated the SimplE model on ten relation-preserving shuffled graphs in which head and tail entities were randomized while relation frequencies were retained **(Fig. 2I)**. Performance collapsed across all metrics on these controls **(Fig. 2J)**, whereas accuracy on the original Ceekr-KG remained high. Relative to shuffled-graph baselines (Hits@1 = ∼14%), the SimplE model achieved a Hits@1 of 97%, corresponding to a 6.9-fold improvement over random structure. Importantly, chance-corrected performance, defined as (H_model_−H_shuffled_)/(1−H_shuffled_) was ∼0.95, indicating that the model captures approximately 95% of the maximum achievable improvement beyond chance. These findings demonstrate that predictive power derives from non-random semantic content rather than graph topology alone.

**Figure 2:**
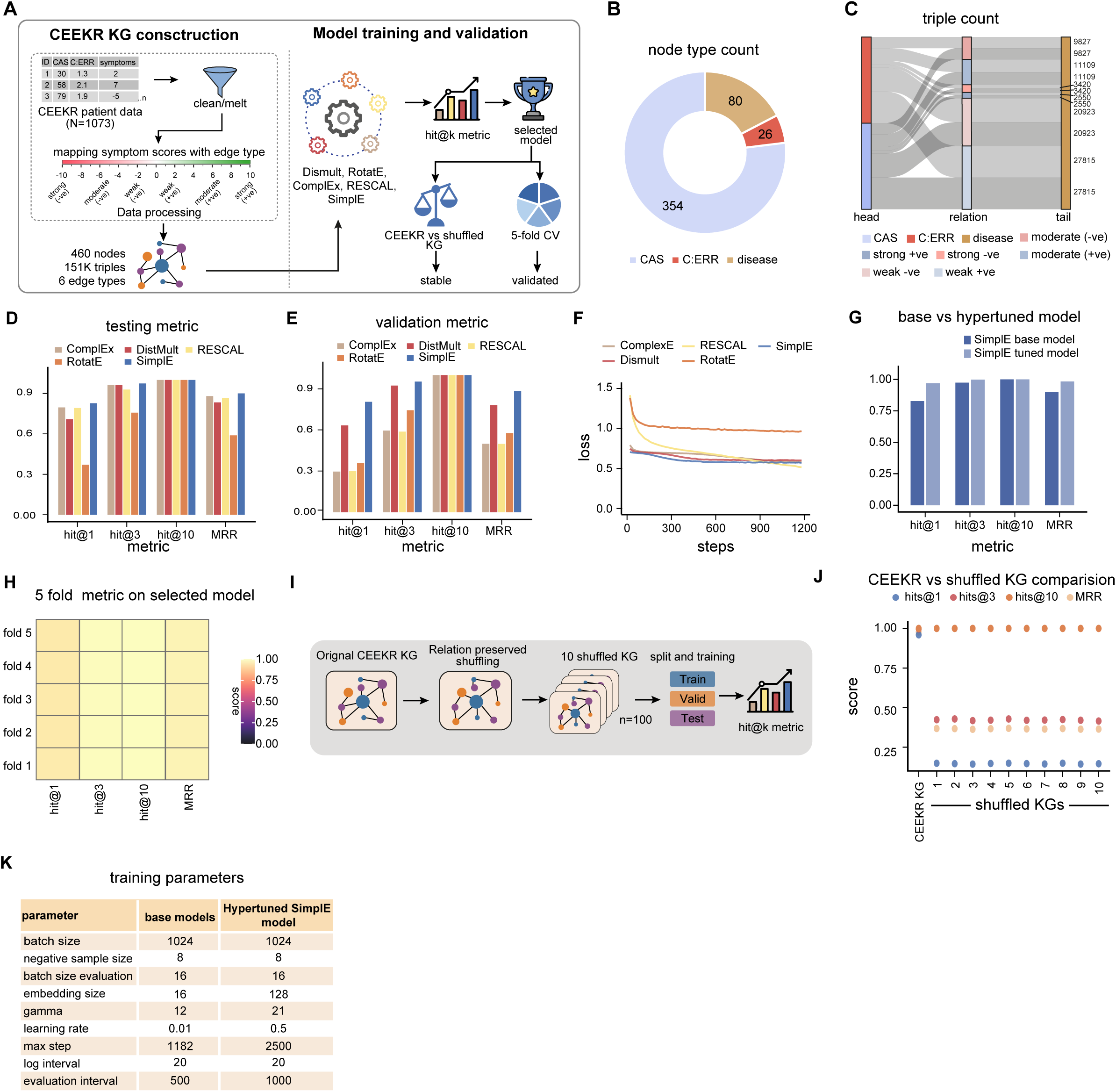
Construction, validation, and predictive performance of Ceekr□KG. **(A)** Schematic of Ceekr□KG construction pipeline. Triples were generated to construct the knowledge graph comprising 460 nodes and approximately 151,288 triples across six edge types. Multiple knowledge graph embedding models (DistMult, RotatE, ComplEx, RESCAL, SimplE) were trained and evaluated using hit@k metrics. Model stability assessed using relation-preserving shuffled KGs, and final selected model validated using 5-fold cross-validation. **(B)** Node type composition of Ceekr□KG. Donut chart showing distribution of 354 discrete CAS levels, 26 C:ERR bins, and 80 clinical symptom nodes, highlighting relative contribution of each entity type. **(C)** Triple composition across head–relation–tail categories. Sankey diagram illustrating distribution of triples across head, relation, and tail types, summarizing how CAS, C:ERR, disease, and symptom polarity categories contribute to overall KG structure. **(D,E)** Base model performance on test set (D) and validation set (E), respectively. Bar plots comparing link prediction performance across embedding models using Hits@1, Hits@3, Hits@10, and mean reciprocal rank (MRR). SimplE achieves highest scores across all metrics on both test and validation sets: test set Hits@1 = 82%, Hits@3 = 97%, Hits@10 = 100%, MRR = 0.90. **(F)** Training loss convergence across models. Loss curves for all base models plotted across optimization steps (0-1,200 steps). SimplE exhibits fastest and smoothest convergence among all models. **(G)** Performance comparison between baseline and hypertuned SimplE models.The hypertuned SimplE model consistently outperforms the baseline across HITS@1, HITS@3, and MRR, while both achieve perfect HITS@10. This demonstrates the effectiveness of hyperparameter optimization in improving link prediction performance. **(H)** Five-fold cross-validation performance of selected SimplE model. Heatmap showing Hits@1, Hits@3, Hits@10, and MRR scores across five folds. Performance remains highly consistent: Hits@1 range 95-96%, Hits@3 range 99%, MRR range ∼97%, confirming robustness and generalizability. **(I)** Schematic of relation-preserving shuffled KG generation and evaluation pipeline. Ten shuffled KGs generated by random reassignment of head and tail entities while maintaining relation frequencies. Each shuffled KG split into training, validation, and test sets and evaluated using hit@k metrics. **(J)** Performance comparison between original Ceekr□KG and ten relation-preserving shuffled KGs. Dot plot comparing link prediction performance using Hits@1, Hits@3, Hits@10, and MRR. Consistently higher performance of original Ceekr□KG indicates predictive accuracy arises from meaningful semantic structure rather than graph statistics alone. **(K)** The table summarizes the key training parameters used for the baseline SimplE model and the hypertuned SimplE (HPT) model. Hyperparameter optimization resulted in increased batch size (1024), embedding dimension (128), gamma (21), learning rate (0.5), and maximum training steps (2500), while maintaining the same evaluation batch size (16) and log interval (20). The evaluation interval was adjusted from 500 to 1000 steps. These modifications were implemented to improve convergence and overall link prediction performance.

### Diagnostic benchmarking of CAS against gold-standard scales via integration with the Clinical Knowledge Graph

Having established Ceekr-KG as a semantically grounded predictive framework, we next benchmarked the Cognitive Attention Score (CAS) against established clinical scales, the Hamilton Anxiety Rating Scale (HAM-A)^[3]^, the General Anxiety Disorder-7 (GAD-7) ^[5]^, and the Hamilton Depression Rating Scale (HAM-D) ^[6]^, using the refined version of the publicly available Clinical Knowledge Graph (CKG)^[17]^, which comprises over 20 million nodes and 1 billion relationships curated from experimental data, public databases, and the biomedical literature (refer to materials and methods). We integrated Ceekr-KG nodes, including CAS, the Cognition-to-Emotional-Response Ratio (C:ERR), and associated symptom entities, into CKG to generate an augmented graph CKG-CEEKR-KG **(Fig. 3A)**. The resulting graph combined Ceekr-derived entities with genes, proteins, diseases, metabolites, tissues, mutations, and ontology terms **(Fig. 3B)** and exhibited diverse relational semantics across node types, as summarized by edge-type composition and chord-diagram analysis **(Fig. 3C)**. We next trained a SimplE embedding model on CKG-CEEKR using optimized hyperparameters (batch size 1,024, negative sampling size 8, embedding dimension 16, learning rate 0.01; **Fig. 3E**). Training showed stable loss convergence **(Fig. 3F)**, and the model achieved strong link-prediction performance on the test set (Hits@1 =85%, Hits@3 = 92%, Hits@10 = 100%, mean reciprocal rank = 0.89; **Fig. 3D**). We next benchmarked anxiety-related inference by encoding questionnaire items from CAS, HAM-A, and GAD-7 as knowledge-graph triples and quantifying their scores with predefined anxiety outcome nodes. To assess whether Ceekr-KG contributes information beyond existing public resources, we compared triple prediction scores obtained using the baseline CKG **(Supplementary Fig. 5A-I)** with those derived from the integrated CKG-Ceekr-KG graph. Incorporation of Ceekr-KG nodes resulted in a systematic increase (more towards zero) in prediction scores across all scales. For anxiety-related outcomes, triple scores for HAM-A, GAD-7, and the Ceekr questionnaire were significantly higher in the integrated graph **(Fig. 3G)**. For cognitive and depressive outcomes, the effect was even more pronounced: HAM-D, the Ceekr questionnaire, and Ceekr-derived nodes all showed marked upward shifts in score distributions following Ceekr integration **(Fig. 3H)**. These results indicate that Ceekr-KG encodes clinically relevant semantic content not captured by existing public biomedical graphs. Finally, ranking symptoms by mean prediction score across all edge types and C:ERR levels identified aging, cognitive decline, asthma, irregular menstrual cycles and nervousness among the top 10 **(Fig. 3I)**. Analysis by edge type highlighted a clear semantic divide, symptoms generally performed more robustly in (+ve) edge types, with cognitive decline serving as an exemplar of stability, while the (-ve) edge categories frequently induced lower medians and increased score dispersion for symptoms like nervousness. The heatmap analysis **(Fig. 3J)** reveals that symptom-disease associations are highly sensitive to disease stage, with irregular menstrual cycles and palpitations displaying maximum prediction scores at specific C:ERR thresholds.

**Figure 3:**
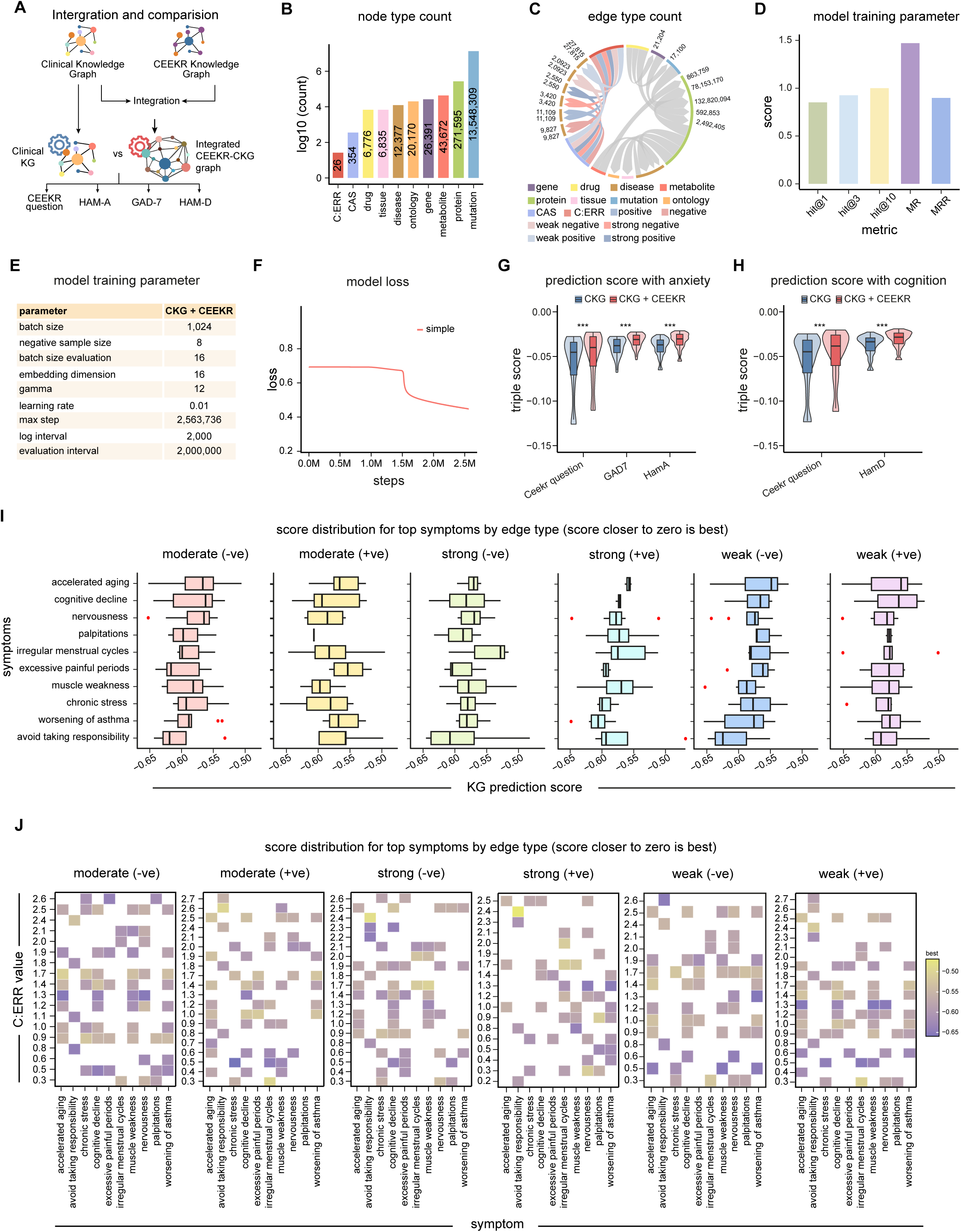
Diagnostic benchmarking of CAS against gold□standard scales via integration with the Clinical Knowledge Graph. **(A)** Schematic illustrating integration of Ceekr□KG with Clinical Knowledge Graph (CKG). Clinical assessment scales (HAM□A, GAD□7, HAM□D) and Ceekr questionnaire data were incorporated to generate an integrated CKG-CEEKR graph used for downstream modeling and comparison. **(B)** Node type composition of the integrated CKG-CEEKR knowledge graph. Bar plot showing log-scaled counts of different node types: C:ERR bins, CAS levels, diseases, genes, proteins, metabolites, tissues, mutations, and ontology terms, highlighting heterogeneous biological and clinical representation. **(C)** Edge type composition of the integrated knowledge graph. Chord diagram summarizing distribution of edge (relation) types, including positive and negative associations with varying strengths, illustrating how different entity types connect through diverse relational semantics. **(D)** Model performance across evaluation metrics. Bar plot showing link prediction performance of SimplE model on integrated CKG-CEEKR graph: Hits@1 = 85%, Hits@3 = 92.4%, Hits@10 = 100%, mean rank (MR) = 1.47, mean reciprocal rank (MRR) = 0.89. **(E)** Training hyperparameters for the integrated CKG-CEEKR embedding model. Table listing key hyperparameters: batch size 1,024, negative sampling size 8, learning rate 0.01, training duration 2.5 million steps. **(F)** Model training loss convergence. Training loss curve of SimplE model plotted across optimization steps, demonstrating stable convergence throughout training. **(G)** Distribution of triple prediction scores for anxiety-related outcomes across knowledge graph variants. Violin plots overlaid with boxplots show distribution of triple scores for anxiety prediction tasks (HAM□A, GAD[7, Ceekr Questionnaire) evaluated using two knowledge graph configurations: CKG (baseline, blue) and CKG-CEEKR (augmented with Ceekr-derived nodes, red). Violin widths represent kernel density estimates; embedded boxplots indicate median and interquartile range. Y-axis represents triple scores (negative values indicate lower plausibility). Statistical significance throughout: Wilcoxon rank-sum test for pairwise comparisons; *p < 0.05, **p < 0.01, ***p < 0.001, ****p < 0.0001; ns, not significant. **(H)** Distribution of triple prediction scores for cognitive impairment and depression-related outcomes. Violin/box plots for HAM□D and Ceekr Questionnaire evaluated using CKG (blue) and CKG-CEEKR (red) configurations. **(I)** Boxplots showing distribution of knowledge-graph prediction scores for top 10 symptoms (identified by overall mean score across all edge types), stratified by relationship (edge) type. Prediction scores closer to zero indicate stronger model confidence. **(J)** Heatmap showing best prediction score for each symptom across C:ERR disease score levels, faceted by edge type. Tile colors represent the maximum prediction score observed for a given symptom-C:ERR combination; darker colors indicate stronger associations. Clear gradients across C:ERR levels indicate certain symptoms become more predictive at specific disease severities rather than uniformly across the entire range. Differences between edge types suggest distinct relational pathways encode severity-dependent information.

### Longitudinal improvement of CAS across three therapeutic interventions

The Ceekr breathing/attention exercise-based therapeutic programmes: AMS (a multimodal wellness intervention), 2M (8-week), and 3M (12-week), operationalize the yogic psychology-derived framework as scalable clinical interventions. To test whether the Cognitive Attention Score (CAS) is sensitive to these therapeutic programmes, we analyzed longitudinal trajectories from participants enrolled in these programmes. **(Fig. 4A)** From the initial matrix comprising approximately 1,840 individuals with CAS measurements, we applied two inclusion criteria: (i) at least two CAS assessments and (ii) a baseline CAS < 64, indicating at least moderate impairment. This yielded 167 individuals in the AMS cohort, 50 in the 2M cohort, and 32 in the 3M cohort, for a total of 249 participants **(Fig. 4B, F, J)**.. For each participant, the first measurement was defined as baseline, and subsequent measurements as follow-up assessments. In the AMS cohort, paired analysis of baseline versus final assessment measurement showed a robust improvement, with mean CAS scores increasing significantly (mean difference = +11.45, 95% CI [8.68, 14.21], paired t-test p < 0.001 **(Fig. 4C)**. The baseline median lay below 60, with a substantial fraction of individuals scoring near 30, whereas after programme engagement the interquartile range shifted upward and the median exceeded 60 **(Fig. 4D)**. Stratification into seven clinical bands (Very Poor: 0-32.5, Poor: 33-44, Medium: 44.5-55, High Medium: 55.5-63.5, Very Good: 64-70, High: 71–88.5, Best: ≥89) revealed pronounced migration out of high-risk categories. “Very Poor” decreased from 40 to 13 and “Poor” from 12 to 0, while higher-function bands increased from near-zero at baseline to 37 (“Very Good”), and 16 (“Best”). The transition matrix confirmed predominantly upward or stable trajectories, with limited downward movement among 167 individuals, 36 remained in “High Medium,” 20 moved from “High Medium” to “Very Good,” and 10 progressed directly from “High Medium” to “Best,” with the densest transitions concentrated in the middle-to-upper bands **(Fig. 4E)**. In the 2M (8 weeks) cohort, paired analysis likewise showed significant improvement, with median CAS increasing from approximately 58 at baseline to nearly 60 at final measurement (paired t-test, p = 0.00256; **Fig. 4G**), indicating that the change was statistically significant. Band analysis showed risk reduction **(Fig. 4H)** “Very Poor” decreased from 10 to 4, and the previously empty “Very Good” and “Best” categories together contained 16 individuals at the final time point. The transition matrix indicated that most participants improved **(Fig. 4I)**, including 10 individuals who moved from “High Medium” to “Very Good” or “Best,” and 6 who progressed from “Very Poor” to “Medium.” In the 3M (12 weeks) cohort, paired analysis revealed highly significant improvement, with median CAS increasing from approximately 58 to approximately 65 (paired t-test, p < 0.001; **Fig. 4K**). The broader follow-up distribution relative to baseline indicates diversification of positive outcomes. Band analysis showed complete elimination of severe-risk categories **(Fig. 4L),** the “Poor” and “Medium” bands (2 and 7 individuals at baseline) were empty at the final measurement. In contrast, no participants initially occupied the “Very Good”, “High” or “Best” bands, approximately 25% reached these categories by the end of the final measurement. The largest fraction migrated from “High Medium” to “Very Good” and “High.” The transition matrix showed no regression **(Fig. 4M)**, with 3 individuals moving from “High Medium” to “Very Good” and “best” and several achieving multi-band gains from “Medium” to “High,” and no entries in lower-right cells indicating downward transitions.

**Figure 4:**
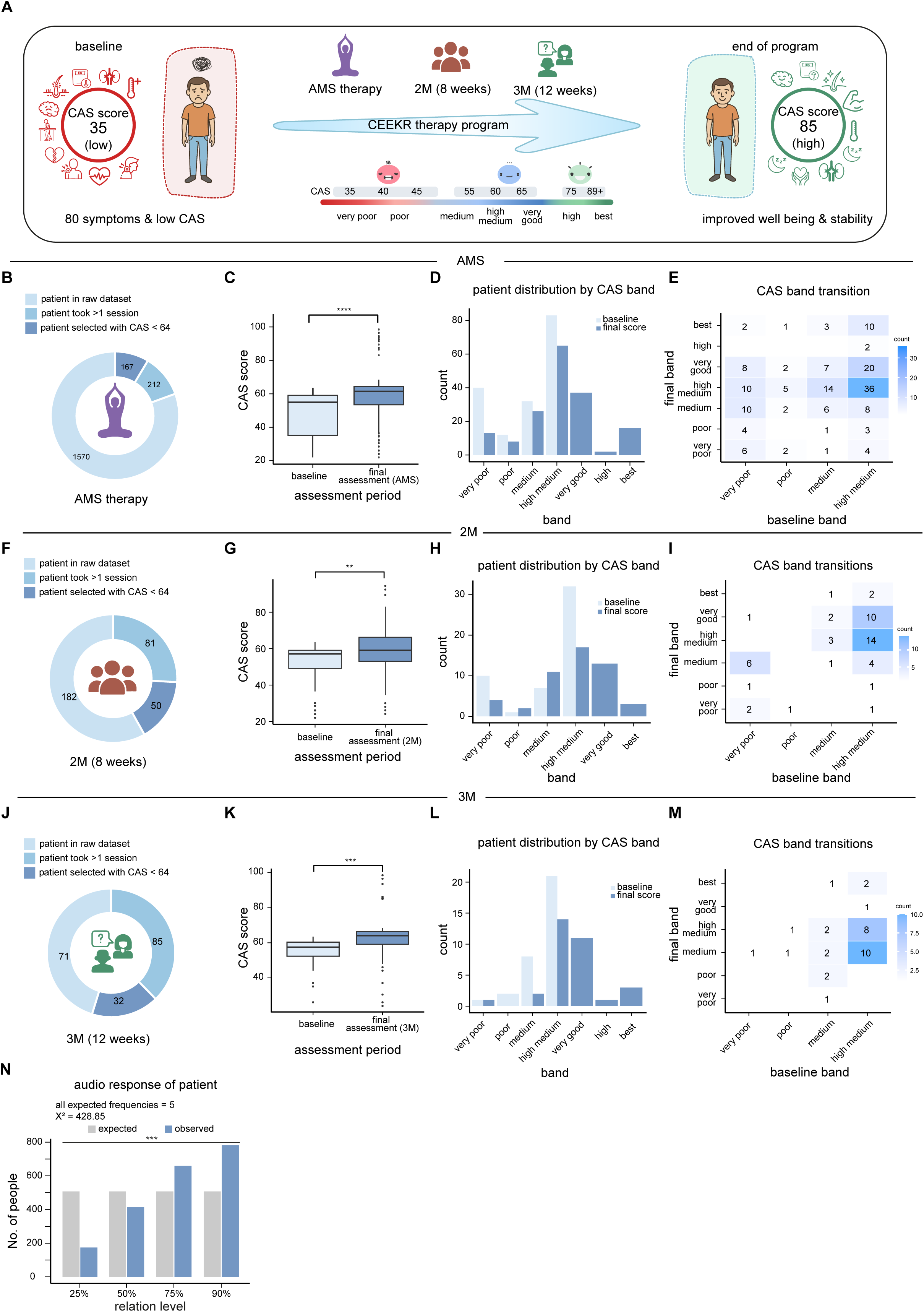
Longitudinal improvement of CAS across three therapeutic interventions. **(A)** Schematic of study design and patient flow. **(B,F,J)** Donut charts illustrating patient selection workflow for AMS therapy (B), 2M (8 weeks) therapy (F), and 3M (12 weeks) coaching (J) cohorts. Subsequent segments indicate patients who completed more than one therapy session and those meeting inclusion criterion of baseline CAS < 64. Final cohort sizes: AMS n = 167, 2M n = 50, 3M n = 32. **(C,G,K)** Paired boxplots comparing baseline versus final assessment CAS scores for AMS (C), 2M (G), and 3M (K) cohorts. Boxes represent median and IQR; whiskers denote range; lines connect paired observations. Statistical significance calculated using paired t-test. AMS: mean increase +11.45 points. 2M: median increase from 57 to 60 (p = 0.00256). 3M: median increase from 59 to 62 (p < 0.001). **(D,H,L)** Patient distribution by CAS band at baseline (light bars) and final assessment score (dark bars) for AMS (D), 2M (H), and 3M (L) cohorts. CAS bands: Very Poor (0–32.5), Poor (33–44), Medium (44.5–55), High Medium (55.5–63.5), Very Good (64–70), High (71–88.5), Best (≥89). **(E,I,M)** Transition matrices showing individual patient movement from baseline band (x-axis) to last final assessment band (y-axis) for AMS (E), 2M (I), and 3M (M) cohorts. Numbers within squares represent patient counts; darker blue indicates higher counts. **(N)** Chi-square goodness-of-fit analysis of patient-reported relation to patient self reporting. Observed frequencies of patient responses (blue bars) compared against expected frequencies assuming uniform distribution across relation levels (25%, 50%, 75%, 90%; grey bars). All assumptions satisfied: independence of observations, mutually exclusive categories, and expected cell frequencies >5. Observed distribution differs significantly from uniform (χ² = 428.85, df = 3, p < 0.001). Higher relation levels (75% and 90%) overrepresented; lower levels (25% and 50%) underrepresented, indicating a non-random pattern of patient-reported engagement. Statistical significance throughout: *p < 0.05, **p < 0.01, ***p < 0.001, ****p < 0.0001. See **Supplementary Tables 5-7** for complete cohort characteristics and additional statistical details.

To assess user-perceived validity of CAS-based interpretations of cognitive, emotional, and physiological states, we collected self-reported feedback from 2,029 participants using a four-level ordinal scale (25%, 50%, 75%, 90%). A chi-square goodness-of-fit test, meeting all assumptions (all expected counts ≥ 5), showed a significant deviation from a uniform distribution (χ² ≈ 428.85, d.f. = 3, p < 0.001; **Fig. 4N**). The 90% category was strongly overrepresented (observed = 781 vs. expected = 507.25), whereas the 25% category was markedly underrepresented (observed = 174), indicating a non-random distribution skewed toward high perceived agreement with the intervention. Across all three programmes, CAS exhibited consistent and statistically significant improvement, with band-transition analyses showing substantial migration from severe impairment to healthy and optimal functional ranges. Notably, the most intensive programme (3M) achieved complete elimination of severe-risk categories without evidence of regression. Together, these results establish CAS as a clinically sensitive longitudinal outcome measure capable of capturing meaningful improvement across different intervention intensities.

## Discussion

This work proposes a knowledge-graph-based framework that operationalizes a yogic psychology-derived conceptual model into two quantitative digital phenotypes, the Cognitive Attention Score (CAS) and the Cognition-to-Emotional-Response Ratio (C:ERR), and demonstrates their utility for mental health assessment, benchmarking, and longitudinal monitoring. Rather than reiterating the empirical findings, we focus here on the conceptual implications, methodological strengths, and limitations of the approach, and on how this framework positions itself relative to existing questionnaire-based and biomedical assessment paradigms. A central strength of this study is the explicit semantic formalization of a phenomenological mental health framework into a machine-interpretable knowledge graph ^[17,24]^. Traditional psychiatric questionnaires, including HAM-A ^[3]^, HAM-D ^[4]^, and GAD-7 ^[5]^, are highly valuable clinically but are structurally limited; they are episodic, construct-siloed, and largely detached from lifestyle, behavioral, and physiological context ^[25,26]^. By contrast, Ceekr’s design embeds cognition, emotion, symptoms, and lifestyle factors into a unified representational space, enabling joint reasoning across domains. This is not merely an implementation detail; it reframes mental health assessment from a set of disconnected scores into a structured state space amenable to computational inference and integration with external biomedical knowledge ^[27,28]^. A second strength is the use of knowledge graphs as a first-class modeling substrate rather than as a passive data container ^[29,30]^. The sharp performance drop under relation-preserving randomization indicates that prediction is driven by semantic structure rather than graph topology, addressing a common criticism of graph-based methods in biomedicine ^[31]^. This supports the interpretation that Ceekr-KG encodes clinically meaningful constraints and relationships, rather than merely correlations induced by network connectivity. Third, the integration with the Clinical Knowledge Graph (CKG) ^[17]^ highlights a practical and often underexplored advantage of semantic representations: bidirectional enrichment. Not only do Ceekr-derived entities gain context from a large biomedical resource, but the addition of Ceekr-KG also improves inference for established scales. This suggests that the Ceekr framework contributes non-redundant, clinically relevant knowledge that is absent from current public resources, and that legacy scales and novel digital phenotypes need not compete; they can co-evolve within a shared semantic ecosystem ^[32,33]^. Finally, the longitudinal analyses underscore a pragmatic strength; CAS functions not only as a screening indicator but also as a sensitive outcome measure capable of capturing graded improvement across different therapeutic intensities. Many existing questionnaires were not designed for dense, real-world longitudinal tracking, whereas the band-based transitions and non-random engagement patterns observed here suggest that CAS is well-suited for continuous monitoring and intervention feedback loops.

Several limitations should be noted. First, although the framework is semantically rich, its primary inputs are self-reported and therefore subject to reporting bias and contextual effects, as with most questionnaire-based scales, motivating future integration with objective signals such as wearable, sleep, or physiological measures. Second, while the cohorts are longitudinal and substantial, they are not population-representative and the interventions were not randomized controlled trials; thus, the present analyses demonstrate sensitivity to change rather than causal treatment effects. Third, the knowledge graph is curated and model-driven, reflecting both data and principled design choices; although robustness tests argue against trivial overfitting, alternative ontological formulations and expanded vocabularies should be explored to assess generalizability. Finally, while integration with the Clinical Knowledge Graph enables systematic diagnostic alignment and large-scale link prediction, the present results support the potential utility of CAS in clinical decision-making. Further prospective studies, including direct cross-comparison of patient cohorts using standard clinical parameters alongside CAS scores, will be important to more precisely define and validate this role.

This work does not argue for replacing established scales such as HAM-A, HAM-D, or GAD-7. Instead, it suggests a complementary paradigm, traditional scales excel at standardized, interpretable snapshots, whereas Ceekr’s metrics operate as context-aware, semantically integrated state variables that can bridge symptoms, lifestyle, and biomedical knowledge ^[17,27,28]^. The observed improvements when Ceekr-KG is merged into CKG illustrate how such a hybrid ecosystem can outperform any single scale in isolation. In this sense, Ceekr functions not merely as a new questionnaire but as an integrative translation layer linking phenomenology, clinical practice, and computational medicine. More broadly, this study highlights a methodological advance in how such conceptual frameworks can be operationalized within data-driven clinical research. Ancient medical and contemplative traditions encode sophisticated systems-level views of mind-body interaction, but they have remained largely disconnected from evidence-based, data-driven medicine due to the lack of formal representations. Knowledge graphs offer a scalable path to co-register historical conceptual frameworks with modern clinical, molecular, and behavioral data without forcing premature reductionism ^[29,30]^. The Ceekr-KG/CKG integration provides a proof of principle for such a synthesis.

## Materials and Methods

### User Data Collection and Consent

All data were collected through the Ceekr digital wellness platform (https://www.ceekr.com/home) or its mobile applications, from users voluntarily enrolled in the AMS, 2M (8-week), or 3M (12-week) programs. Prior to enrollment, all users completed a single-stage digital consent process within the application and intended use for research and product improvement. Users confirmed agreement by selecting a check-box, with consent timestamps automatically recorded. Collected data included Ceekr questionnaire and username and email address. All personally identifiable information was removed prior to analysis, with each user assigned a unique pseudonymized identifier; the mapping to personal information is stored separately in an encrypted, access-controlled database. Data handling procedures comply with applicable data protection regulations, including GDPR and HIPAA. As this study involved analysis of de-identified data collected through routine platform operations and did not constitute human subjects research under federal regulations, Institutional Review Board (IRB) review was not required.

### Data Collection and Curation

The foundational knowledge matrix underlying this study was constructed through systematic manual translation of classical Yogic psychological frameworks into a structured computational format. Drawing from primary texts and traditional commentaries, a panel of subject matter experts, including practitioners collaboratively mapped qualitative descriptions of mental states, cognitive processes, and their relationships to physical and emotional symptoms onto a quantifiable framework. This translation process resulted in a comprehensive matrix encoding the directional relationships between 80 clinically observed symptoms and the latent constructs of cognitive-attentional function and emotional dysregulation. From this knowledge matrix, two composite metrics were derived: the Cognitive Attention Score (CAS) and the Cognition-to-Emotional Response Ratio (C:ERR). The mathematical formulations underlying CAS and C:ERR are proprietary to Ceekr Concepts and represent the operationalization of yogic psychological principles into quantitative computational constructs. The final curated matrix, provided in its entirety as **Supplementary Table 1**, encodes the quantitative relationships between 354 discrete CAS levels, 26 C:ERR bins, and 80 clinical symptoms across six relationship types capturing both the polarity (incremental or decremental) and strength of association. This matrix served as the primary input for constructing the Ceekr Knowledge Graph (Ceekr-KG), wherein each relationship was instantiated as a subject-predicate-object triple following standardized knowledge graph conventions. The translation from matrix format to graph representation preserved all semantic information while enabling downstream graph-based predictive modeling and inference. Data for the Clinical Knowledge Graph (CKG) was integrated into the Ceekr-KG framework using the official Neo4j database distribution provided by the CKG maintainers<u>(Santos et al. 2022)</u>.

### Knowledge Graph Embedding (KGE) Training

Vector representations of entities and relations were learned using the Deep Graph Library (DGL, version 1.1.2)^[34]^, and DGL-KE (version 0.1.0)^[35]^. Five KGE models (RotatE, SimplE, DisMult, RESCAL, ComplEx) were systematically evaluated. The triples were split into training (80%), validation (10%), and test (10%) sets. Base models were trained with an embedding dimension of 16, a batch size of 1024, and a margin loss function. The optimal architectures were identified as SimplE for Ceekr-KG and CKG-CEEKR-KG **(Supplementary Table 4).**

### Computational Infrastructure

Computational training and assessment were conducted on a high-performance Linux workstation running Ubuntu 22.04 LTS and CUDA 12.0. The system utilized NVIDIA RTX 5000 Ada Generation GPUs with 32 GB of VRAM, providing the necessary memory and parallel processing power to manage a knowledge graph training and analysis.

### Statistical Analysis

Statistical analyses were performed using R (v 4.2.3), primarily utilizing the tidyverse suite for data manipulation and the ggplot2, ggpubr, and patchwork packages for high-quality visualization. Continuous variables, specifically CAS and C:ERR, were characterized through descriptive statistics including means, standard deviations, and ranges, while normality was rigorously assessed using Shapiro-Wilk tests supplemented by visual inspection of Q-Q plots and Gaussian kernel density estimates. Raw symptom data were binned into an ordinal scale of six severity levels ranging from Strong Negative (-10 to -7) to Strong Positive (7 to 10), and their prevalence was evaluated through frequency analysis of the top 12 symptoms. To quantify the relationship between CAS and C:ERR, both Pearson’s r and Spearman’s rho coefficients were calculated. Furthermore, Spearman’s rank correlation was employed to evaluate the monotonic associations between the binned symptom severity levels and continuous outcomes, while a comprehensive Spearman correlation matrix was constructed for the top symptoms to identify inter-symptom patterns. Group-level differences in scores across the severity bins were evaluated using one-way ANOVA **(Supplementary Tables 2,3)**. For the longitudinal AMS, 2M and 3M therapy analysis, a cohort was established by filtering for patients with a baseline CAS < 64 and ≥ 2 observations. Clinical improvement was statistically validated using paired-samples t-tests comparing the first (Baseline) and final (“On Program”) scores for each participant. Finally, categorical shifts in clinical status were evaluated using transition matrices to map movement between CAS performance bands from baseline to study conclusion. The threshold for statistical significance was set at alpha = 0.05 for all tests.

## Data Availability

All data produced in the present work are contained in the manuscript and its supplementary files. Specifically, patient-related metrics and clinical association tables are provided in Supplementary Tables 1-7. The complete source code for the Ceekr-KG platform and the fully trained models are publicly available on Zenodo at https://doi.org/10.5281/zenodo.18595967

## Data and Code Availability

The complete source code for the CEEKR KG platform, alongside the full trained model generated is publicly available on Zenodo at https://doi.org/10.5281/zenodo.18595967.

## Conflict of Interest

The authors declare no competing research or financial interests related to this work. Prof. Gaurav Ahuja previously served as a scientific consultant for Ceekr Concepts Private Limited; however, no active consultancy agreement was in place during the preparation of this manuscript. Ankur Jain, Shivani Modi, and Kaushal Kumar are founders of Ceekr Concepts Private Limited.

## Acknowledgments

The authors gratefully acknowledge the entire team at Ceekr Concepts Private Limited for their valuable discussions and collaborative support. The computational infrastructure and IT facilities provided by Indraprastha Institute of Information Technology Delhi (IIIT-Delhi) are sincerely appreciated. We also thank Saveena Solanki for providing valuable comments and suggestions on the figures.

## Author Contributions

The study was conceived by K.K., A.J., S.M., and G.A. Computational analysis workflows were designed by G.A and A.S. Statistical guidance was provided by G.A. Data compilation was performed by A.J., K.K., S.M., and A.B. Modeling workflows were designed and executed by A.S. Illustrations were drafted by A.S. and G.A. wrote the paper. All authors have read and approved the manuscript.

## Supplementary Figure Legends

**Supplementary Figure 1:**
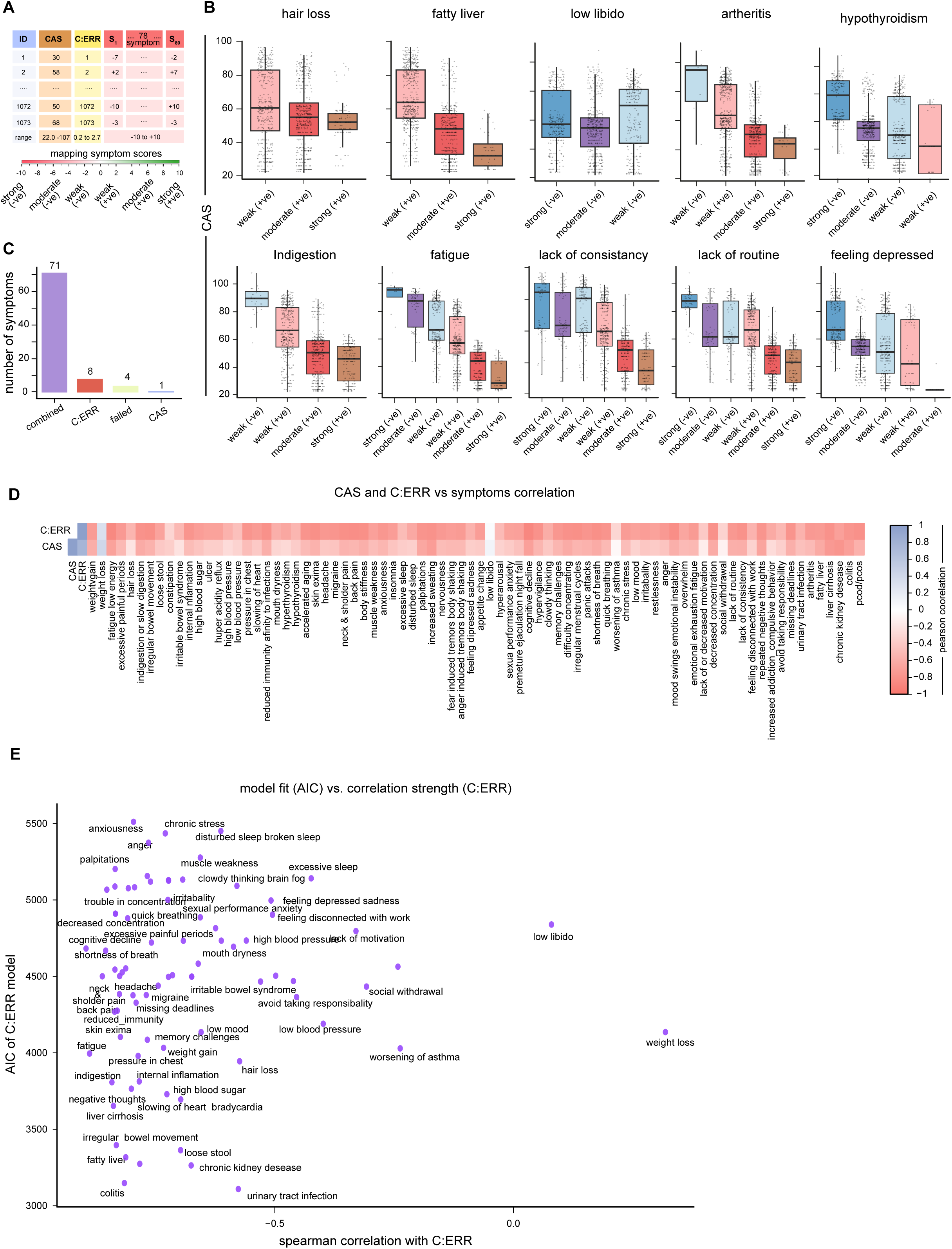
Metrics overview, and bivariate assessment of predictive accuracy. **(A)** Dataset overview. The metric with complete CAS and C:ERR measurements. CAS scores ranged from 22.06 to 107.89, C:ERR values ranged from 0.2 to 2.7, and a total of 80 clinical symptoms were assessed across physical, cognitive, and emotional domains. **(B)** Comparative boxplots of CAS across top clinical indicators. Multi-panel display showing the distribution of CAS scores across severity levels (strong negative to strong positive) for the most frequently reported symptoms. Symptoms, including fatigue, indigestion, and lack of routine, exhibit consistent downward trends with increasing severity, validating the heatmap results in Fig. 1E. **(C)** Frequency of the best predictor model across symptoms. Bar chart showing number of symptoms (of 80 total) for which each ordinal regression model achieved the lowest AIC: combined CAS + C:ERR model (n = 71), C:ERR only (n = 8), CAS only (n = 1), and failed fits (n = 0). **(D)** Heatmap showing Pearson correlation coefficients between symptom severity scores and the predictors CAS and C:ERR. Each column represents an individual symptom; each row corresponds to one predictor. Color intensity indicates direction and magnitude of correlation, with warmer tones (red/brown) representing stronger negative correlations and cooler tones (blue) representing stronger positive correlations. Correlation values range from −1 to 1. C:ERR consistently exhibits stronger negative correlations across most symptoms compared to CAS, confirming its superior sensitivity to symptomatic distress. **(H)** Bivariate assessment of predictive accuracy and association strength. Scatter plot correlating Spearman rank correlation coefficient with C:ERR (x-axis) against Akaike Information Criterion (AIC) for C:ERR-only model (y-axis). Each point represents a distinct clinical symptom. Lower AIC values (y-axis) signify more robust and parsimonious model fit.

**Supplementary Figure 2:**
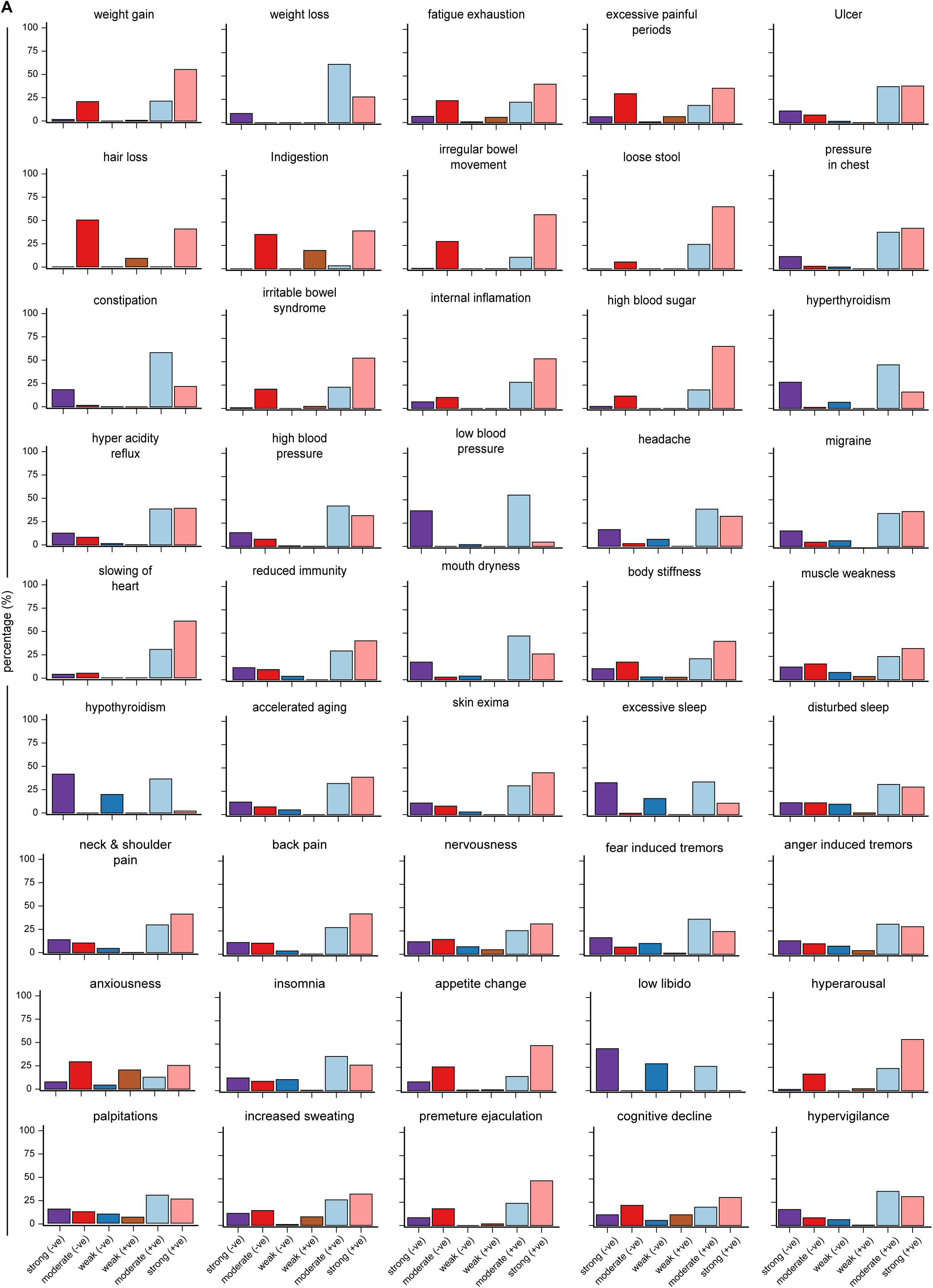
Percentage distribution of severity levels for the indicated symptoms. Each panel represents one symptom, with bars indicating the proportion of observations within each binned severity category. Symptoms are displayed in batches of twelve panels per page for readability. Severity categories derived from binned symptom scores; percentages calculated relative to total non-missing observations for each symptom. Most symptoms show skewed distributions with higher frequencies concentrated in lower-severity categories, while subsets exhibit broader or more uniform distributions, indicating greater variability in symptom expression. Heterogeneity observed across symptoms highlights differences in prevalence and severity patterns, providing essential context for ordinal modeling and predictor analyses.

**Supplementary Figure 3:**
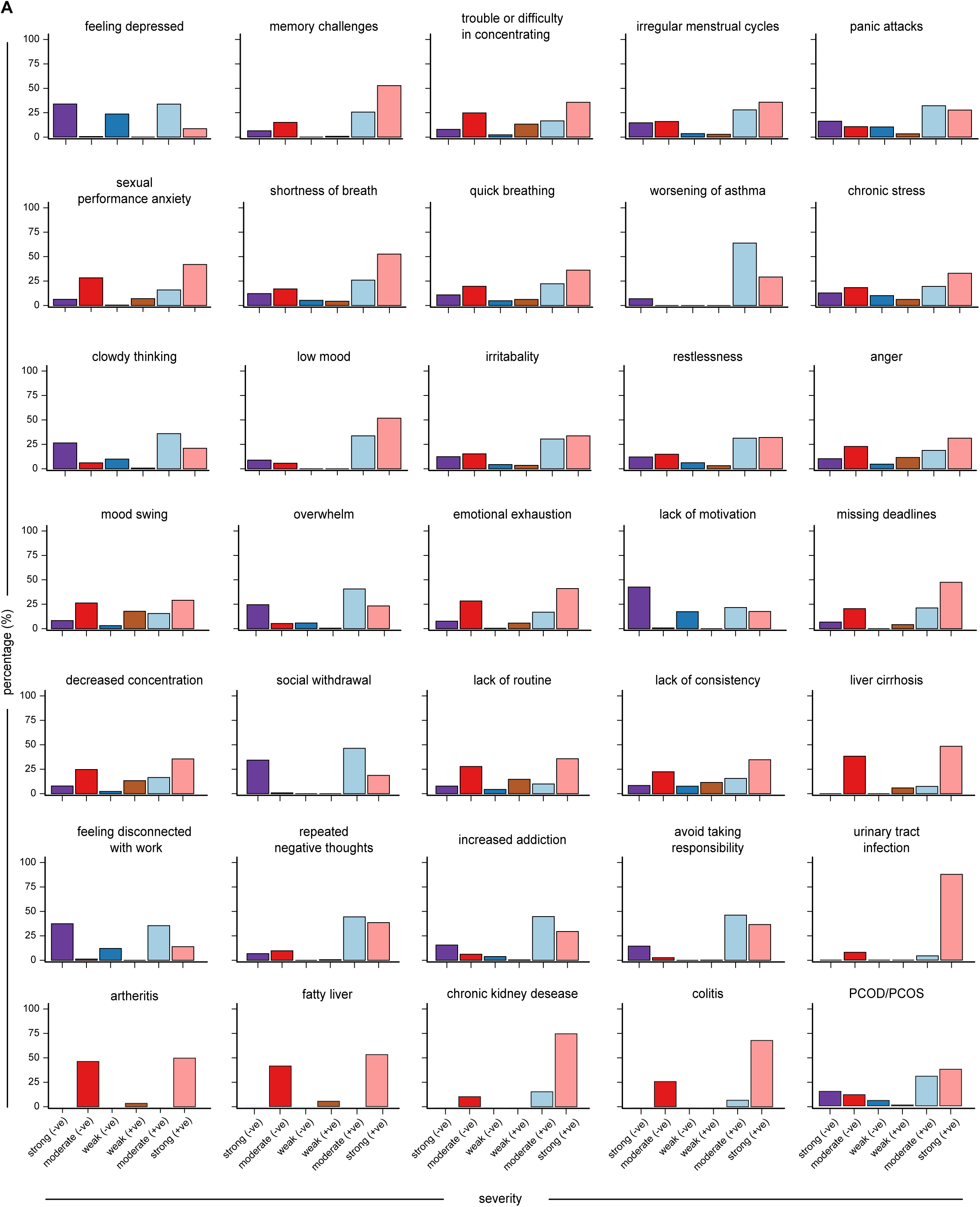
Percentage distribution of severity levels for the indicated symptoms. Continuation of severity distribution plots for the remaining 40 clinical symptoms. Format and interpretation as in Supplementary Figure 2. Together, Supplementary Figures 2 and 3 provide complete documentation of severity distributions for all 80 symptoms assessed in the study cohort.

**Supplementary Figure 4:**
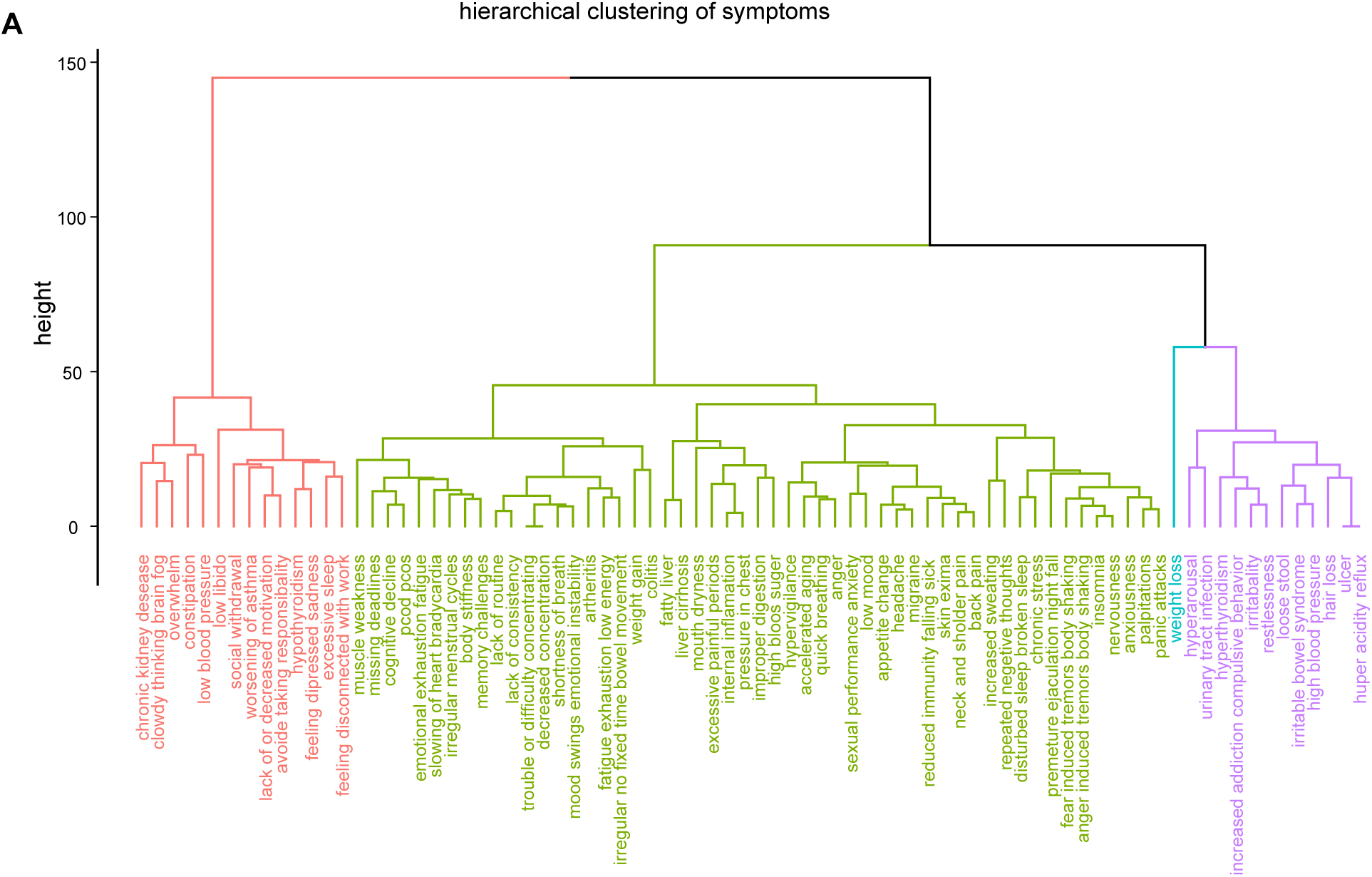
Complete correlation heatmap and hierarchical clustering of symptoms. **(A)** Hierarchical clustering of symptoms based on severity profiles. Dendrogram showing clustering of symptoms into distinct groups with similar severity patterns across individuals, suggesting shared underlying pathophysiological mechanisms. Clusters reveal groupings of related symptoms (e.g., metabolic symptoms, cardiovascular symptoms, cognitive symptoms) that may inform future mechanistic investigations and targeted intervention strategies.

**Supplementary Figure 5:**
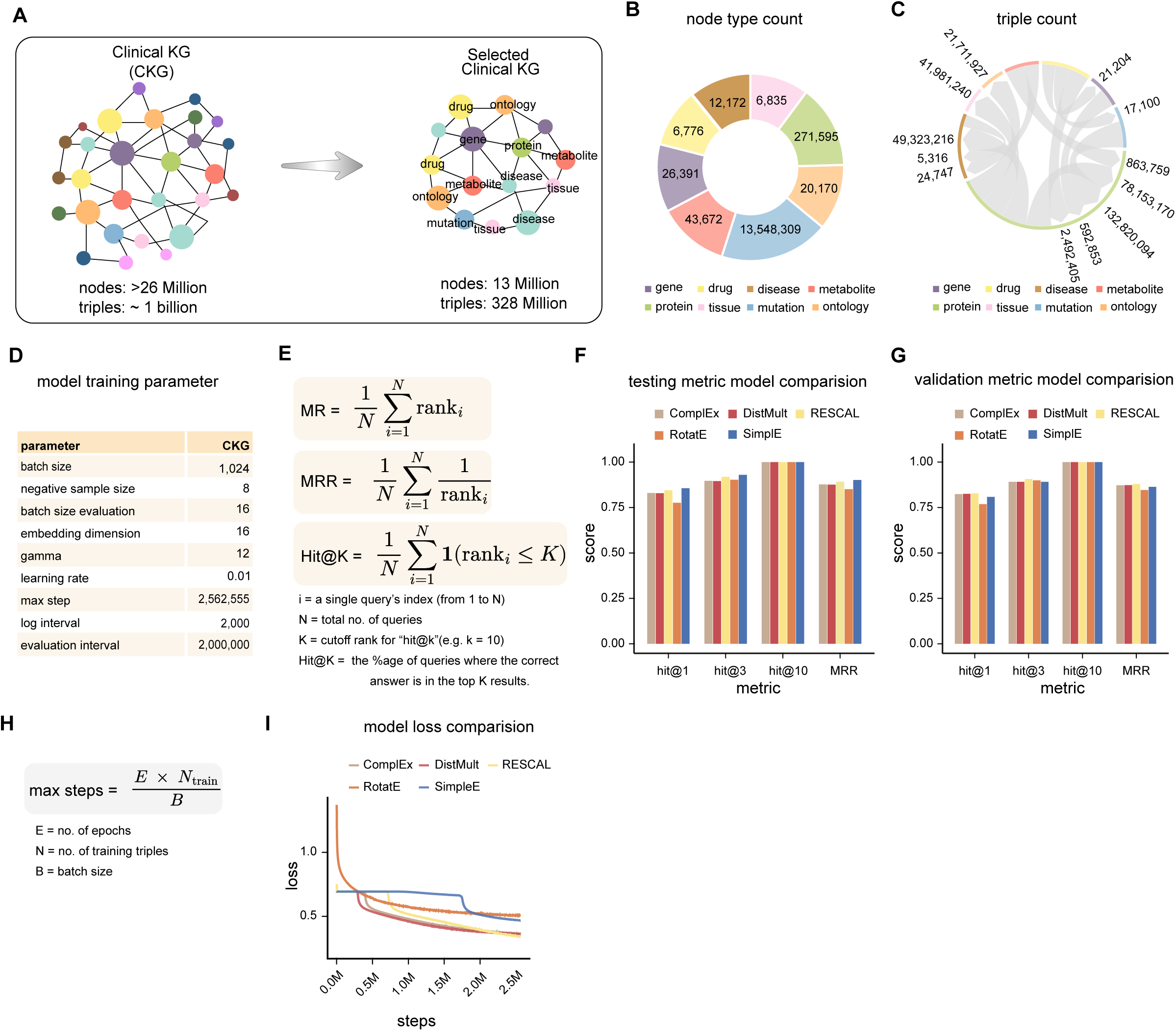
Construction, composition, and training of the Clinical Knowledge Graph subgraph. **(A)** Clinical knowledge graph selection and construction. Schematic illustrating transition from full Clinical Knowledge Graph (CKG; ∼20 million nodes, ∼700 million edges) to selected subgraph used for model training. A subset of biologically and clinically relevant entity types—including gene, drug, disease, protein, metabolite, tissue, mutation, and ontology terms, was retained, resulting in a reduced but semantically focused KG for efficient training and evaluation. **(B)** Node type composition of selected clinical knowledge graph. Donut chart showing distribution of node types in selected CKG subgraph: genes (largest proportion), proteins, drugs, diseases, metabolites, tissues, mutations, and ontology terms, highlighting heterogeneous biological and clinical representation. **(C)** Edge type composition of selected clinical knowledge graph. Chord diagram summarizing the distribution of edge (relation) types between different entity categories. Edge widths represent the number of triples associated with each relation type, illustrating connectivity structure and density of relationships across biomedical entity classes. **(D)** Model training parameters. Table listing key hyperparameters used for training knowledge graph embedding models on CKG subgraph: batch size 1,024, negative sampling size 8, learning rate 0.01, evaluation intervals, and training duration. **(E)** Definition of evaluation metrics. Mathematical definitions of ranking-based evaluation metrics used for link prediction: mean rank (MR), mean reciprocal rank (MRR), and Hits@K. Lower MR and higher MRR and Hits@K indicate better model performance. **(F)** Model performance comparison on the test set. Bar plots comparing link prediction performance of multiple KG embedding models (ComplEx, DistMult, RESCAL, RotatE, SimplE) on CKG subgraph test set using Hits@1, Hits@3, Hits@10, and MRR. SimplE demonstrates superior performance across all metrics. **(G)** Model performance comparison on the validation set. Validation set performance of the same embedding models evaluated using Hits@1, Hits@3, Hits@10, and MRR, demonstrating consistency between validation and test results, with SimplE maintaining optimal performance. **(H)** Training step calculation. Formula illustrating computation of total training steps based on number of epochs, training triples, and batch size: total steps = (epochs × training triples) / batch size. **(I)** Training loss convergence across models. Loss curves for different KG embedding models plotted over training steps (0-2.5 million steps), showing convergence behavior and relative optimization dynamics. SimplE exhibits fastest and most stable convergence among all models.

## Supplementary Tables

**Supplementary Table 1.**

The table provides a full nomenclature for all 80 clinical symptoms included in the Ceekr knowledge matrix and analyzed throughout the study. Symptoms span physical domains (e.g., cardiovascular, gastrointestinal, musculoskeletal), cognitive domains (e.g., attention, memory, executive function), and emotional domains (e.g., mood, anxiety, stress responses).

**Supplementary Table 2.**

Comprehensive statistical analysis of symptom associations with CAS and C:ERR. For each symptom, descriptive statistics (mean, SD), one-way ANOVA results (F-statistic and p-value), and correlation coefficients (with corresponding p-values) are reported. Multiple testing correction was applied using the Benjamini–Hochberg false discovery rate (FDR) method where indicated.

**Supplementary Table 3.**

This table summarizes the distributional characteristics of the primary continuous variables (CAS and C:ERR), including measures of central tendency, dispersion, range, skewness, and kurtosis. These statistics were used to assess normality assumptions and inform the selection of appropriate statistical tests.

**Supplementary Table 4.**

It presents the complete comparative evaluation of embedding models across the CEEKR KG, CKG KG, and combined knowledge graphs. Standard link prediction metrics (HITS@K, MR, MRR) are reported for validation and test sets to ensure transparency and reproducibility of model performance.

**Supplementary Table 5.**

The table presents baseline demographic characteristics, clinical profiles, and CAS band distributions for individuals enrolled in the AMS multi-modal wellness programme. The data includes participant ID, CAS score, report date, and programme start date, capturing repeated assessments from baseline to final recorded measurement. Only participants with at least two CAS assessments and a baseline CAS < 64 were included in the analytical cohorts.

**Supplementary Table 6.**

The table presents baseline characteristics for individuals enrolled in an 2M (8-week) programme. The table includes participant ID, CAS score, report date, and programme start date, capturing repeated assessments from baseline to final recorded measurement. Only participants with at least two CAS assessments and a baseline CAS < 64 were included in the analytical cohorts.

**Supplementary Table 7.**

The table presents baseline characteristics for individuals enrolled in a 3M (12-week) coaching programme, The data includes participant ID, CAS score, report date, and programme start date, capturing repeated assessments from baseline to final recorded measurement.

## Notes

### Competing Interest Statement

The authors have declared no competing interest.

### Funding Statement

This research was supported by Ceekr Concept Private Limited

### Author Declarations

The Institutional Review Board (IRB) of Ceekr Concept Private Limited waived ethical approval for this work because it involved the analysis of de-identified data collected through routine platform operations.

